# Variation in social contact levels and changes during the COVID-19 pandemic by occupation and socio-economic status in east Zimbabwe

**DOI:** 10.64898/2026.10.04.26364721

**Authors:** Sophie Ayling, Constance Nyamukapa, Rufurwokuda Maswera, Louisa Moorhouse, Rebekah Morris, Brian Moyo, Alessia Melegaro, Owen Mugurungi, Hannah Fry, Sarah Wise, Guy Harling, Simon Gregson

## Abstract

**Background:** COVID-19 movement restrictions reduced social contact worldwide, curbing viral spread but with unequal economic consequences where social safety nets are limited. Little evidence exists on how these impacts varied by occupation, wealth and age in resource-constrained settings. This paper examined pre-pandemic contact and lockdown-related change.

**Methods:** The paper documents analysis of a population-representative survey (N=8,499) in Manicaland, Zimbabwe, during April-May 2021, capturing typical weekday contacts before and during the first COVID-19 lockdown. Negative binomial regression models assessed pre-lockdown contact counts by socio-economic characteristics, overall and by place (home, school, work, transport, entertainment); linear regression assessed predictors of lockdown declines.

**Results:** Pre-COVID-19, respondents reported a median of 12 daily contacts (interquartile range [IQR] 4-47), with higher counts among manual labourers (incidence rate ratio [IRR] 2.21, 95% confidence interval [CI] 2.00, 2.44), retail and public sector workers (IRR 2.33, 95%CI 2.09, 2.60) and students (IRR 2.59, 95%CI 2.38, 2.82), males, wealthier respondents, under-30s, and urban and agricultural estate residents. Counts were highest in schools. During lockdown, contacts fell across all subgroups to a median of 8 (IQR 4-28). Declines were greatest among those with most pre-COVID-19 contacts: urban (mean −17.5, 95%CI −19.53, −15.37) and agricultural estate residents (−18.1, 95%CI −19.99, − 16.13), students and teachers (−17.7, 95%CI −19.82, −15.59), males and 30-44 year olds. Drops were greatest in schools and on transport.

**Conclusion:** Reductions in contact under blanket lockdown varied markedly between subgroups and places in Manicaland, Zimbabwe. Selectively targeting high-contact groups might have achieved similar transmission reductions while limiting economic damage, holding lessons for similar settings.

## Introduction

Data on the numbers and nature of population-level social contacts are essential for informing infectious disease models, which in turn feed into outbreak prediction and response (Buckee, Noor and Sattenspiel, 2021) This pipeline has been instrumental in combating both airborne and human-to-human contact diseases, including tuberculosis, Respiratory Syncytial Virus (RSV), Severe Acute Respiratory Syndrome (SARS), measles and influenza (Frank *et al*., 1983; CDC, 2005, 2016, 2018, 2020; NFID, 2022). A key aspect of social contact patterns is that non-random mixing has important implications for disease dynamics – both in terms of initial speed of spread and ultimate epidemic size (Milwid *et al*., 2019). It is now increasingly possible to model heterogeneous mixing patterns though data-informed, granular models, such as Agent Based Models, thereby increasing the utility of having such data.

Most research on how social contact patterns are non-random has focused on age-based mixing, largely in High Income Countries (HICs) where employment is predominantly formal (Hoang *et al*., 2019a; Leung *et al*., 2023). There have been a minority of similar studies in Low and Middle Income Countries (LMICs). In sub-Saharan Africa (SSA) these include work in Kenya, Malawi, South Africa, Uganda and Zimbabwe (Melegaro *et al*., 2017; le Polain de Waroux *et al*., 2018; Quaife, van Zandvoort, *et al*., 2020; Del Fava *et al*., 2021; McCreesh *et al*., 2021; Thindwa *et al*., 2021). This pattern of data collection creates two inter-related potential problems. First, LMICs – where the majority of infectious disease morbidity and mortality arises – must rely on ‘synthetic’ contact matrices projected from other settings (Prem, Cook and Jit, 2017). These estimates may be inaccurate if mixing patterns vary by cultural and wealth context. For instance, a systematic review found that while in HICs average daily contact counts decrease with age, this is not the case in LMICs (Mousa *et al*., 2021). Second, the focus on age-structured mixing matrices fails to account for other forms of stratification, which may have practical policy relevance, such as where contacts occur and how they may vary by occupation or other socio-demographic characteristics. Again, this can be exacerbated when HIC and LMIC patterns differ. For example, LMIC homes see higher contact numbers compared to HICs due to LMIC homes more often being large and intergenerational. LMIC social contact surveys do indeed show contact patterns characterised by: (i) greater age assortativity (individuals interact more often with similar-aged peers, especially school-aged children and young adults); and (ii) greater intergenerational mixing (Del Fava *et al*., 2021). Both of these issues underscore the importance of increasing knowledge of LMIC contact patterns.

During the COVID-19 global pandemic that was declared on 30^th^ January 2020 by the World Health Organization, social contact data were vital for understanding who was at greatest risk of becoming infected with and transmitting SARS-CoV-2 and the impact of government policies on these risks. However, despite an increase in social contact studies during the pandemic, the skew towards HICs data remained – only two of 12 early social contact pattern studies were done in SSA (Quaife, Van Zandvoort, *et al*., 2020)(McCreesh *et al*., 2021), and two more in China (Liu *et al*., 2021). However, as well as its direct health impact, the COVID-19 pandemic had an economic impact often considered just as severe, particularly in Low- and Middle-Income Countries (LMICs) where strict movement restrictions had severe repercussions for incomes, nutrition, education and earning capacity (Narayan *et al*., 2022). Early in the pandemic, across nine LMICs, a median of 70% of respondents reported declining income with 36% pausing all work, and substantial knock-on impacts for food security (Egger *et al*., 2021) and particularly severe effects in informal urban African settings (Quaife, van Zandvoort, *et al*., 2020). Losses caused by a five-month global school shutdown in 2020 were estimated to have cost a net present value of US$10 trillion (Azevedo *et al*., 2021; Miguel and Mobarak, 2022).

This interconnection of health and economic impacts highlights the importance of examining occupational differences in social contact patterns, and its limited consideration in the context of COVID-19. Some HIC studies of contact patterns during Covid did consider socioeconomic differences in part. For example, the European CoMix studies stratified by wealth class, gender and region in addition to age, but did not analyse whether these factors contributed to differences in contact numbers (Gimma *et al*., 2021; Backer *et al*., 2023). A US study measured workplace contacts, but only within three large US companies, which are unlikely to be representative of employment patterns in LMICs (Kiti *et al*., 2021). Another US study disaggregated contact numbers by occupational status and found that both pre- and post-vaccine roll out, contact rates were highest at work, with significantly higher values for those working in retail, hospitality, food services and transportation (Nelson *et al*., 2022). The two SSA COVID-19 contact studies were conducted in South Africa and Kenya, and again considered socioeconomic factors only partially. In rural South Africa contact counts fell substantially during the first epidemic wave (June-July 2020) from a mean of 7.4 close interaction contacts to 4.1, but while evaluating changes in contacts by age, gender, resident place and household wealth the study did not examine changes by occupation (McCreesh *et al*., 2021). Similarly, in Kenyan informal settlements in Nairobi, researchers found a large reduction of 62-67% in eigen values of contact matrices depending on the pre-COVID-19 matrix used. They also found clear wealth disparities in these contact rates during movement restrictions in May 2020, but this study also did not look at differences by occupation (Quaife, van Zandvoort, *et al*., 2020).

Given the high levels of informal economic activity and strict Covid-19 lockdowns in many LMICs, there is a need for a clearer understanding of how social contacts in LMICs are patterned by occupation and the differential impact of policies to reduce contacts. If blanket restrictions have regressive impacts, it may be worth considering whether more selectively targeted restrictions that consider occupational differences could limit immediate and long-term economic damage at the same time as mitigating health risks, especially where social safety-nets are minimal. While social protection spending increased drastically worldwide in 2020-21, social assistance, social insurance and labour market expenditures were only $28 per capita in SSA, compared to $4,130 in North America, highlighting the limited support LMIC governments could provide (Gentilini *et al*., 2022). Furthermore, the more thinly social assistance is distributed, the less impact it can have to help those in greatest need. A clearer understanding of how broad measures affected behaviour by socioeconomic status can inform design of more targeted measures to maximise targeted assistance (Aiken *et al*., 2023).

Zimbabwe offers a good opportunity to evaluate these questions since about three-quarters of all employment is informal (World Bank, 2023), one of highest reported rates globally but representative for SSA youth (UNDP, 2022). The Zimbabwean government imposed a strict lockdown from March to May 2020, including curfews, prohibition of inter-city movement, closure of all but essential shops and all entertainment venues, and banning of public gatherings (except funerals of 30 people or less) (Chitungo *et al*., 2020; Morris *et al*., 2024). The country subsequently eased restrictions on businesses and intercity travel, but schools were closed until July 2020, and again between January and March 2021 (see Fig S1 & Table S1 for details). These policies are likely to have affected individuals variably depending on their household wealth and occupation.

This paper therefore explored how social contact patterns differed prior to and during COVID-19 lockdowns in eastern Zimbabwe both by occupation – considering formal and informal workers, students and non-workers – household wealth and age. Specifically, this paper identified levels of, and factors associated with, pre-lockdown contact numbers and changes in counts and distributions during lockdown. Such information may be used to inform infectious disease models, in particular agent based or compartmentalised models which hold a comparative advantage for scenario modelling based on context-specific population data. Such scenario modelling could include examining the impact of selective school or workplace-related movement/contact restrictions on the spread of disease within a population. Beyond Zimbabwe, the data may be useful for other contexts with similar occupational breakdowns or composition of small towns and agricultural sites.

## Materials and Methods

### Ethics approval and consent to participate

The Manicaland Survey was approved by the Imperial College Research Ethics Committee (20IC6436) and the Medical Research Council of Zimbabwe (MRCZ/A/2703). This analysis was approved by the University College London Research Ethics Committee (project ID 20527/001). All ethics approvals are available in S1 Text. All participants provided audio-recorded (for telephone interviews) or written (for face-to-face interviews) informed consent prior to participation. Consent for those aged under 18 in the study was provided through written assent. Consent forms are available for review in S2 Text.

### Study Setting and Population

This study used data from the individual questionnaires of round 8 of the Manicaland General Population Open Cohort Survey (“Manicaland Survey”) conducted in the eastern-most province of Zimbabwe and capturing information on wealth characteristics and sexual risk behaviour (Gregson et al., 2017; Rao et al., 2022). The Manicaland Survey is currently conducted in eight sites in four districts representing several location types, ranging from rural (tea estates, subsistence farms, forestry estates) to peri-urban towns to urban Mutare, the provincial capital. These sites represent all main location types in the province of Manicaland except commercial mining and a few border posts; they also capture the five major wealth strata in Manicaland. For round 8, an initial household census was conducted within all eight study sites listed in Table S2. From this census, a two-thirds random sample of households was drawn and all resident men and women aged 15 years and above in these households were eligible for an individual interview. This age-range was chosen to capture older school attendees, working-age adults and the elderly. The Round 8 individual data used in this study were collected in period two of this round, between 24 February to 27 July 2021, for most of which Zimbabwe was in a medium, Level 2 COVID-19 lockdown (Fig S1 and Table S1). Interviews were conducted either over the telephone (for all individual and most household census interviews) or face-to-face (for some household census interviews).

### Data collection

Round 8 of the Manicaland Survey included questions on social contacts. For this survey, the definition of a contact was one commonly used in contact literature (Hoang *et al*., 2019b): ‘an interaction between two individuals – either physical, involving skin-to-skin contact such as a handshake, or non-physical, involving a two-way conversation with three or more words in the physical presence of each other but with no skin-to-skin contact.’ Participants were asked a fixed series of questions pertaining to: (1) the ‘current/most recent lockdown period’; and (2) ‘February 2020 *before* the first COVID-19 lockdown’.

For both timepoints, questions were asked for ‘a typical weekday’ and then for ‘a typical weekend day’ separately for six places: home; work; school (for school goers); public transport; entertainment venues (bars/restaurants/nightclubs/shebeens); and any other place – and four age categories of contacts (<15; 15-44; 45-64; and ≥65). An example phrasing was: ‘How many contacts did you have in a typical [weekday/weekend day] at [place] in the following age-groups?’. The primary measure of interest here was the sum of contacts across all age-groups across all place types on a ‘typical weekday’ and separately analysed the sum of all contacts reported in the place types home, work, school, transport and entertainment types (for the questionnaire section as it appeared to participants see Table S3). Though weekend data was also available, the difference between weekday and weekend samples was not significant, and given that the focus of this study is on occupation-based mixing at work, weekday reported data was retained as the focus.

In the survey, occupation was measured in 16 categories which were aggregated into seven groups with similar characteristics: manual labourers; students or teachers, retail or public service, office workers, informal traders, other informal workers and not employed/homemakers. The site type of their dwelling was divided into five categories (see Table S2). Respondents’ household wealth was measured based on a household asset index that was computed as the proportion of a count of sellable (e.g., radio, TV, bicycles) and non-sellable (e.g., water and electricity supply, toilet facilities, household structure) assets present (Schur *et al*., 2015). Finally, the analysis included individual covariates capturing age categories (15-29, 30-44, 45-64 and 65+) and gender, as well as study site and district.

### Contact counts

Our primary dependent variable for pre-lockdown was the total number of contacts reported per individual respondent across home, school/work transport, entertainment venues or any other place reported per person, on a ‘typical day’ prior to the first COVID-19 lockdown. The same variable was constructed for during lockdown based on a question about a typical day ‘during the current/most recent lockdown period’ (i.e., prior to April-June 2021). An upper bound of 200 was applied to the place-level total number of contacts an individual could report on a weekday, beyond which values were recoded to missings. Separately, the sum of contacts *across places* was winsorized to 200 if values were over that. For place-level contacts, two versions of the aggregates were created at place and population level to correctly manage structural zeros in means where individuals did not visit certain places. Calculating population averages would include everyone and count ‘did not visit a place’ as zero. Meanwhile, place-level averages, i.e., the mean contact count among those who reported visiting a given place was calculated separately as these would be a useful parameter for modelling contact estimates within specific place types. These decisions were based on a thorough review of practices dealing with outliers in past contact literature (Gimma *et al*., 2021; Backer *et al*., 2023) (for more details, see Text S3).

### Covariates

The main focus of this analysis was occupation which in prior studies had been included as a binary covariate (Mousa *et al*., 2021), but not in a disaggregated form, as it is here with 7 categories. Several other socio-demographic covariates have been included. Gender (binary), age (as a continuous variable but categorised into in five subgroups in analysis) and wealth quintiles (as calculated according to an index designed by (Schur *et al*., 2015)) were justified as they had been included as covariates in multiple other contact studies identified in a meta-analysis. Other studies had also tried to examine urban vs. rural distinctions (le Polain de Waroux *et al*., 2018; Del Fava *et al*., 2021) but not disaggregating further to see if there were any differences by site type such as whether the location was an agricultural estate or a commercial centre. Site type, referring to the type of location the respondent was classified as residing in, was therefore also analysed in five subcategories (Table S2).

### Statistical analyses

After pre-processing and cleaning, the research first calculated the median and interquartile range (IQR) of pre-lockdown and lockdown contacts across the population and for subgroups defined by gender, household wealth, age, site type and occupation. Pearson’s *χ*^2^ tests for binary variables and Kruskal-Wallis test for categorical variables were used to test for differences between groups. By place (home, school/work, transport and entertainment) means for those who visited each place were used instead of population level statistics due to the high numbers of structural zeros (i.e., people who did not visit a place, especially during lockdown).

Next the researchers conducted negative binomial regression models (due to overdispersion counts) with household-level random intercepts to account for non-independence between individuals in the same household. These were used to examine associations between pre-lockdown contact counts (overall and by place), and our socio-economic covariates. As a sensitivity analysis, researchers ran an alternative model specification with clustered standard errors and household-level random effects.

Finally to explore changes in contacts during lockdown, the difference in mean contact counts between pre- and post-lockdown introduction were calculated, both overall and for each place. Linear regression was then used to identify predictors of this difference, using the same socio-economic variables as in the rest of the analysis. The marginal differences between pre-lockdown and lockdown counts were then plotted for each group of covariates controlling for all other variables in the model.

## Results

Round 8 of the Manicaland Survey interviewed 8,499 of 9,383 eligible individuals (90.6%) in 3919 households. Due to winsorization rather than exclusion of individuals with over 200 contacts per day, the analytic sample for the main outcome variable remained at 8,499 respondents in both periods (see Text S3 for details of winsorization). The sample was more female than the national population (57.8%) but reflected the wider population’s age with 40.4% of the population aged 15-29 and only 7.5% aged over 65 (Table 1). The plurality of people resided in commercial centres within towns (28.4%), then agricultural estates (22.1%), with almost equal proportions living in roadside trading settlements and urban areas (17.7% and 18.0% respectively), and the lowest proportion (13.8%) living in subsistence farming communities. In terms of occupation, the largest occupational group are unemployed or homemakers (44.7%), followed by 15.7% students or teachers, 12.2% manual labourers and 19.0% being in some kind of informal employment. Office workers form a tiny minority of less than 1% of the population.

**Table 1:**
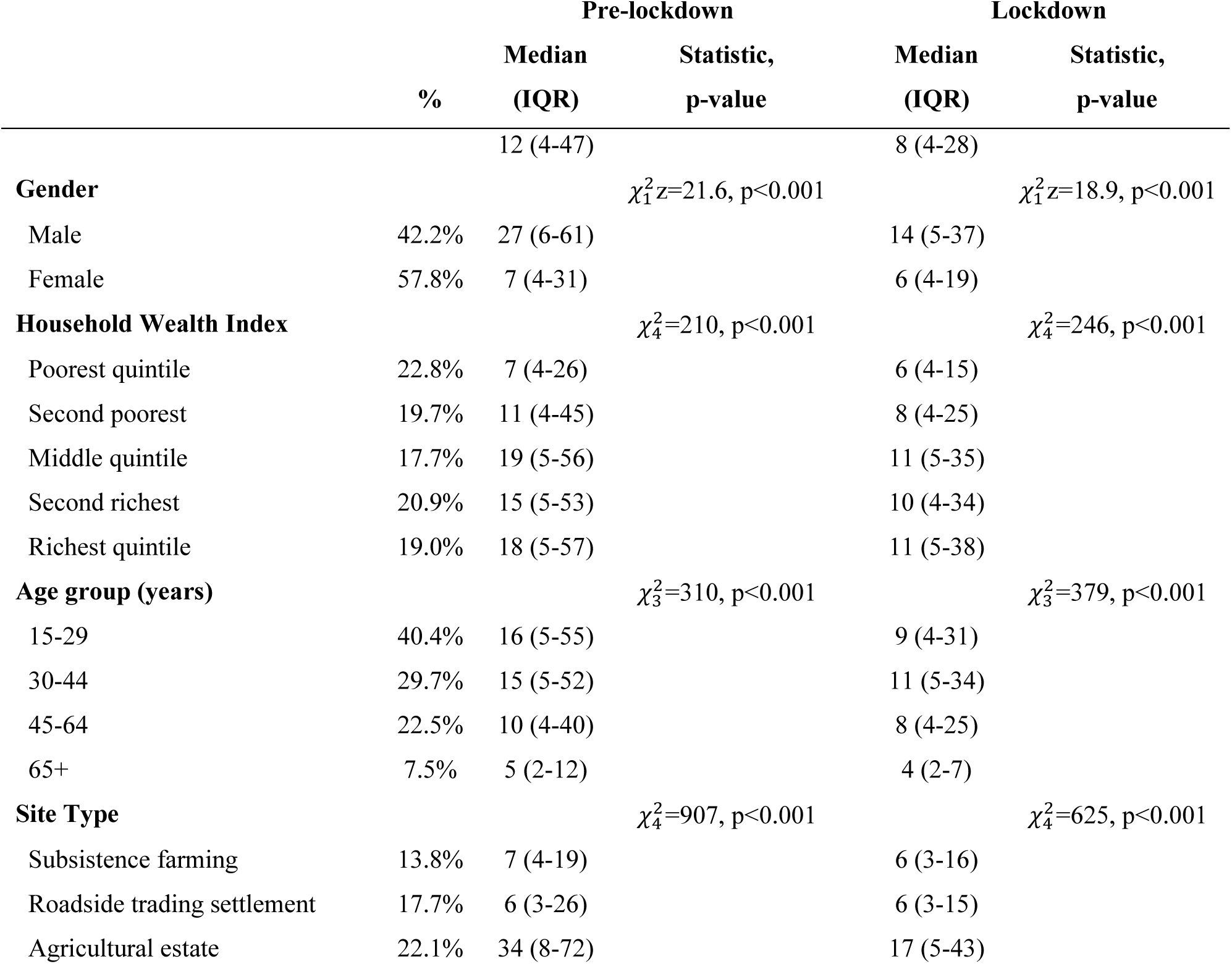
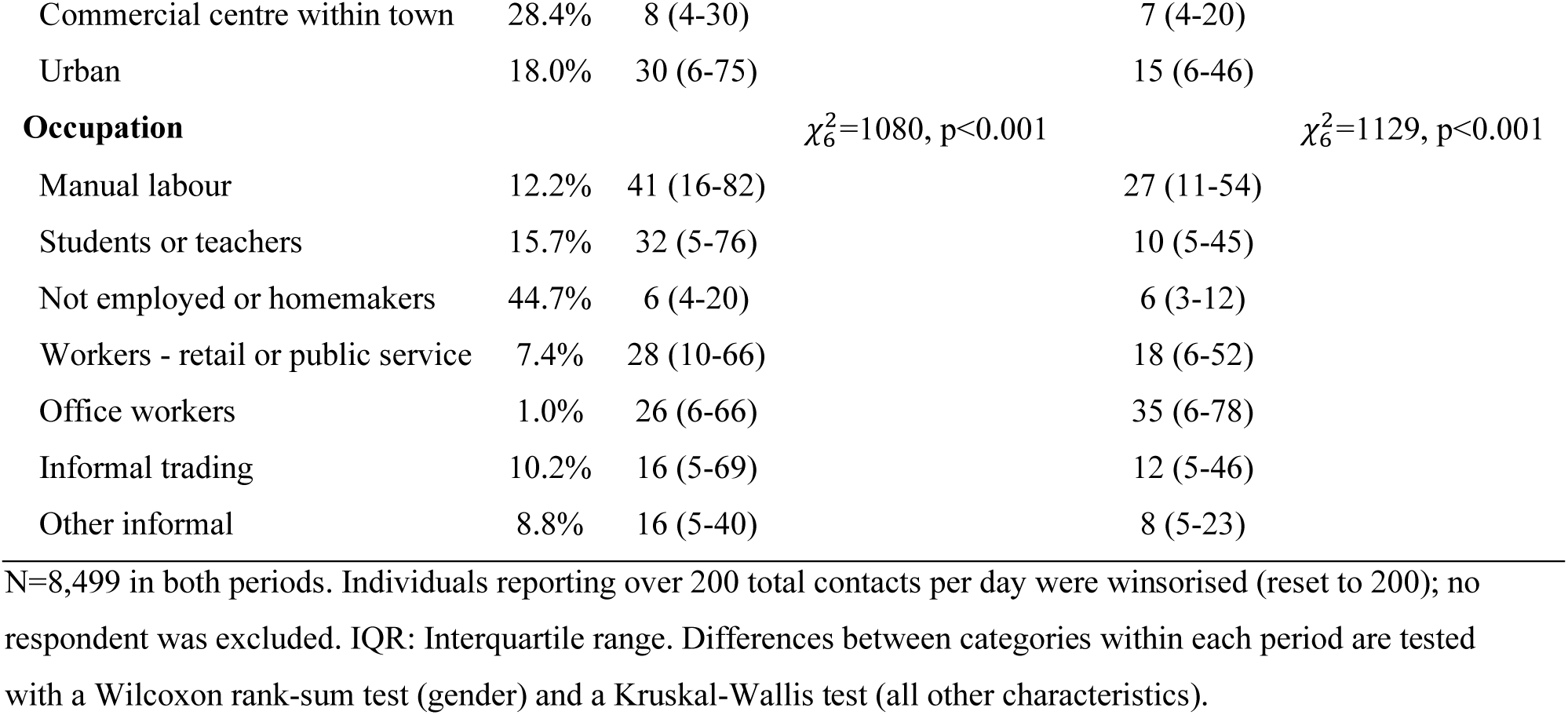
Descriptive statistics for pre-lockdown and lockdown contacts.

Pre-lockdown, a median of 12 (IQR 4-47) daily contacts was reported across the whole population (Figure 1 and Table 1). Reported contacts were highest in manual labourers and students, at intermediate levels for office workers and informal workers, and lowest for the unemployed and homemakers. Reported contact numbers were nearly four times higher for men than for women (male median 27, IQR 6-61; female median 7, IQR 4-31, p<0.001). Contact numbers were significantly lower for the lowest vs. highest wealth quintile, and for those aged over 45 compared to 15-29 year-olds. Urban and agricultural estate residents had significantly more contacts than those in subsistence farm settings.

**Figure 1:**
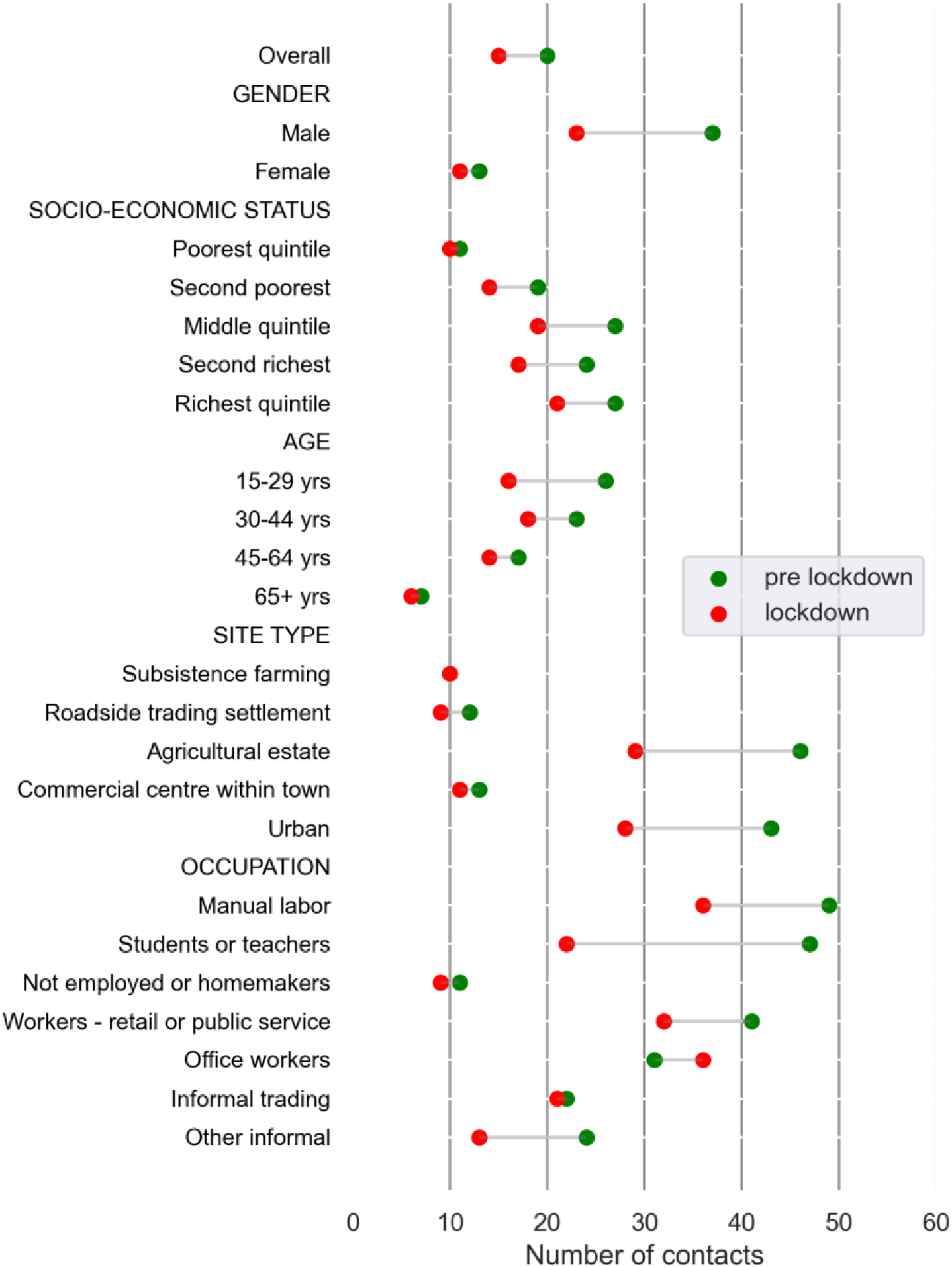
Median contact rates in lockdown and pre-lockdown periods. Participants understood pre-lockdown as pre-February 2020. While for lockdown, this was referenced as the ‘current or most recent lockdown period’ which would have been Jan-Feb 2021 Level 4 measures (Table S1). Source is the R8 Manicaland survey.

When data were separated by place of contact, respondents reported a mean of seven contacts at home which implies a higher than average household size in Manicaland compared to the rest of Zimbabwe, as this was six in the last national census (IPUMS, 2022). Those who visited schools had an average of 46.7 contacts there (Figure 2). The highest average contact numbers (other than at school) for those who visited that place were at work (mean 31.8), then transport (21.0), and then in entertainment venues (16.3). However, means can be skewed by high end outliers and reported place level medians are lower. For school (41), work (13), entertainment (5) and transport (17). Across the population as a whole, taking into account proportion of those visiting each place, mean contacts were still highest at school (28), next at work (12) and on transport (10.3), followed by entertainment venues (2.8) since only a minority of individuals visited entertainment venues. Home contact numbers remained consistent.

**Figure 2:**
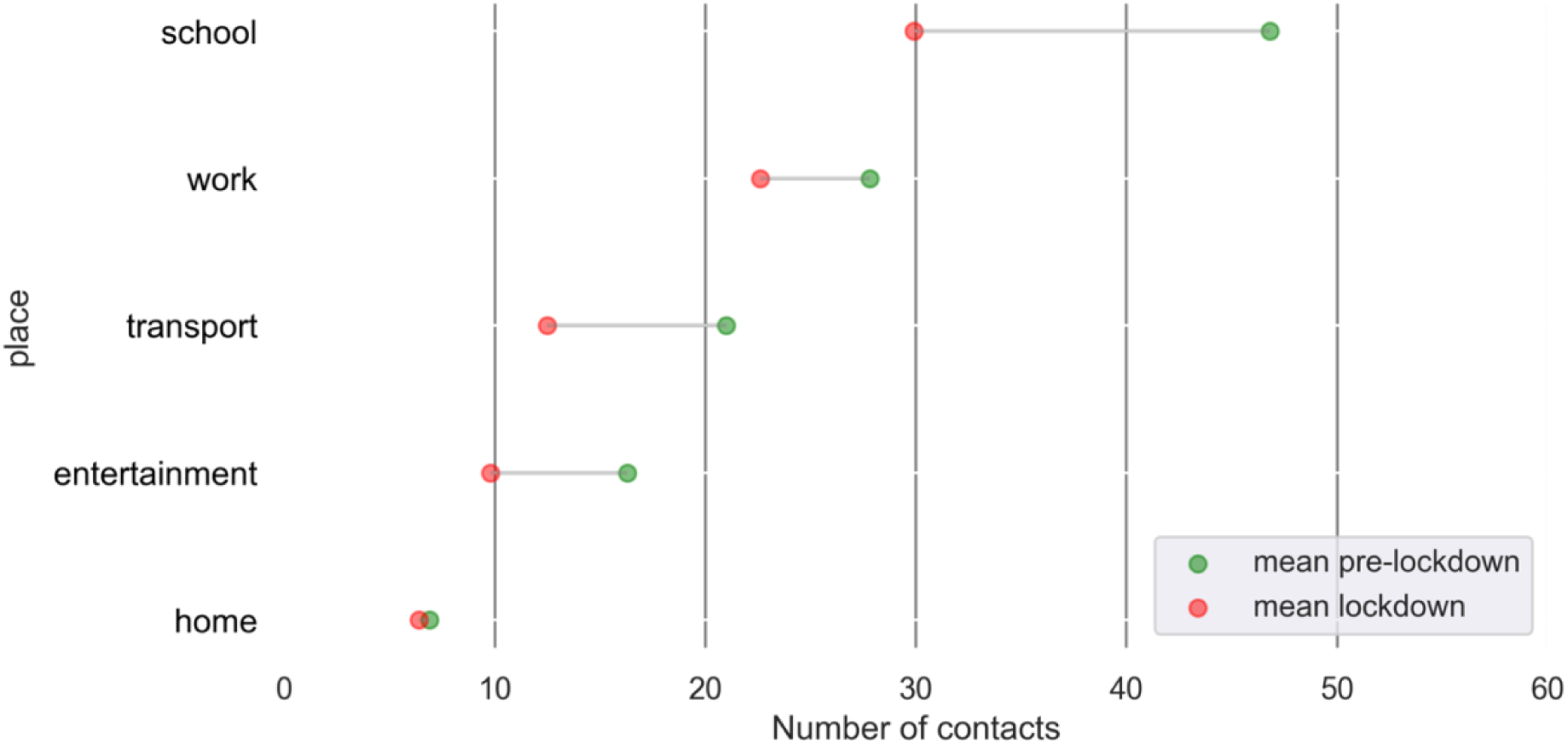
Comparison between mean contact rates in lockdown and pre-lockdown periods among those that visited each contact place type.

In multivariable regression analysis, higher numbers of contacts pre-lockdown were independently predicted by occupation (students or teachers, service workers), living on agricultural estates or in urban areas, being male, and being young (Table 2).

**Table 2:** Multivariable negative binomial regression showing socio-economic factors associated with higher contact numbers overall for individuals on weekdays pre-lockdown with mixed effects.

|  | IRR [95%CI] |
| --- | --- |
| Gender (vs male) |  |
| Female | 0.61 [0.58 ,0.65] |
| Wealth Status (vs poorest) |  |
| Second poorest quintile | 1.07 [0.97 ,1.18] |
| Middle quintile | 1.12 [1.01 ,1.24] |
| Second richest | 1.02 [0.92 ,1.13] |
| Richest quintile | 1.19 [1.07 ,1.32] |
| Age (vs 15-29 years old) |  |
| 30-44 | 1.10 [1.03 ,1.18] |
| 45-64 | 0.83 [0.77 ,0.89] |
| 65+ | 0.57 [0.51 ,0.64] |
| Site Type (vs subsistence farming) |  |
| Roadside trading settlement | 0.87 [0.77 ,0.97] |
| Agricultural estate | 1.92 [1.71 ,2.16] |
| Commercial centre within town | 0.88 [0.79 ,0.98] |
| Urban | 2.08 [1.83 ,2.35] |
| Occupation (vs not employed or homemaker) |  |
| Manual labour | 2.21 [2.00 ,2.44] |
| Students or teachers | 2.59 [2.38 ,2.82] |
| Workers - retail or public service | 2.33 [2.09 ,2.60] |
| Office workers | 2.51 [1.91 ,3.30] |
| Informal trading | 2.07 [1.88 ,2.28] |
| Other informal | 1.76 [1.58 ,1.95] |

IRR: Incidence rate ratio; CI: Confidence interval.

Weekday contacts fell following lockdown, with a marginal reduction of 10.14 contacts (Figure 3, Table S6). This drop was greater for males, younger age-groups and those at school, and in entertainment venues. Only office workers reported an increase in number of contacts, but this change was not statistically significant, reflecting a small number of such workers in the sample. The smallest marginal drop was for workers in retail or public service, and among the unemployed and homemakers.

**Figure 3:**
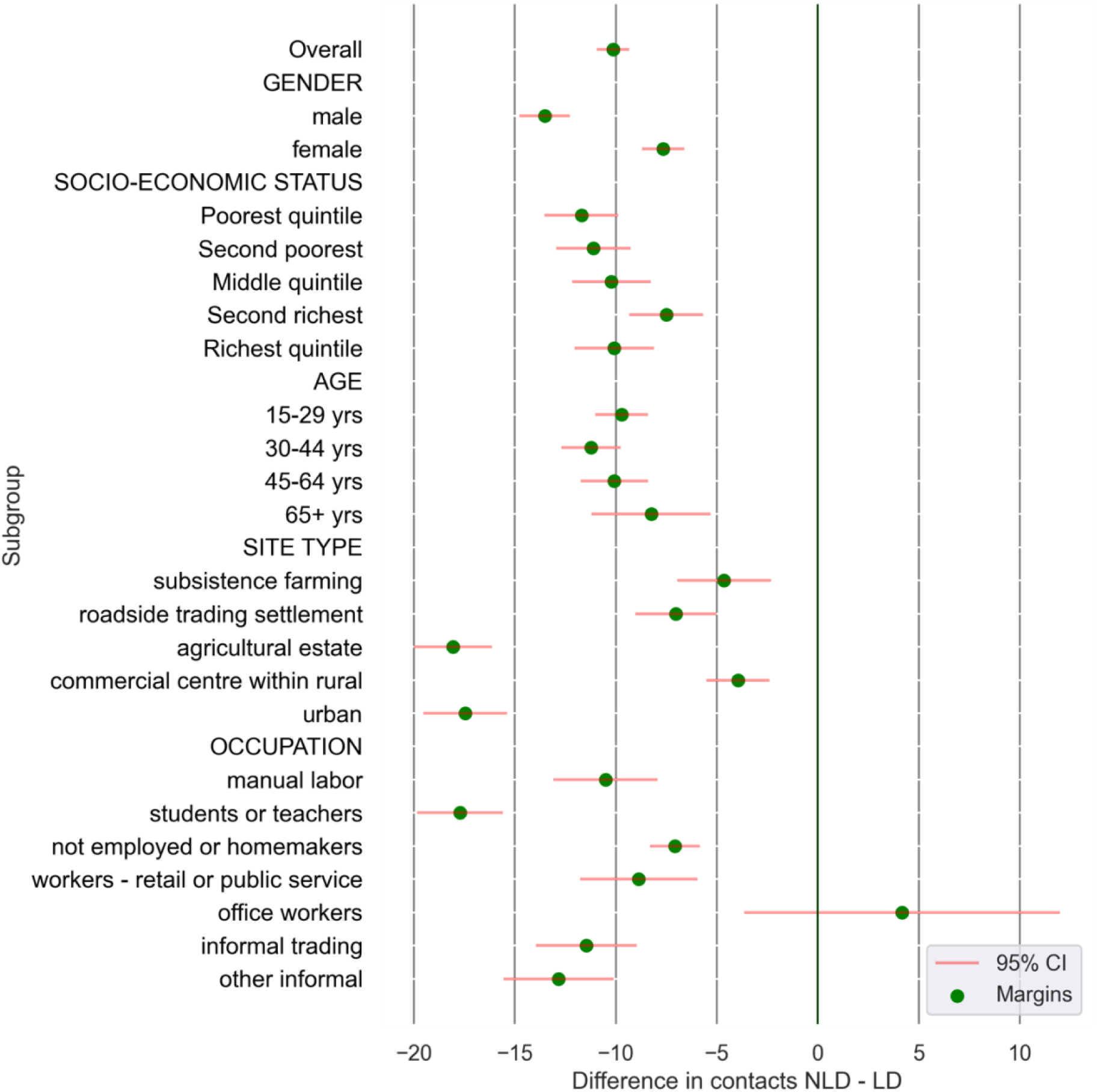
Marginal differences in contact rates from multivariable regression models comparing lockdown and pre-lockdown periods. PLD: Pre-lockdown; LD: Lockdown

In multivariable linear regression, the greatest reductions in contacts were associated with being: a teacher/student; male; and with residents of urban areas and agricultural estates (Table S6 and Table S7). When contacts were separated by location (Table S8), home contacts fell most in agricultural estates and urban centres; work contacts fell most for those aged 30-44 years old, school contacts fell substantially for teachers/students, informal workers and the younger age-groups; transport contacts fell most for informal workers, as well as retail or public service workers; and entertainment contacts fell more for males. Across site types, residents from agricultural estates and urban areas reported the biggest drop. By place, the biggest raw drop in contacts was evident in the number of contacts at school (Table S5). The clustering and mixed effects models do not show substantial differences in coefficients and standard errors (appendix Table S4 for table comparison)

## Discussion

This paper represents one of the few studies on pre-post COVID-19 lockdown social contact numbers with a sample size of over 8000 residents, representative across a mix of site types in a largely rural province of Zimbabwe, in Eastern Africa. This is relevant not just for historical record, but also to inform future modelling, policy and planning with regards to application of movement restrictions in response to an infectious disease outbreak passed through human-to-human contact. It provides data on the occupations that were most vulnerable to SARS-CoV-2 infection and at the greatest risk of spreading the disease during the COVID-19 pandemic in a sub-Saharan African population due to their higher contact rates, and on those that were most impacted by movement restrictions. The study found that among the residents of Zimbabwe’s Manicaland province, individuals’ self-reported social contacts pre-COVID-19 varied substantially by occupation, place of contact and place of residence, as well as by more-studied axes of age, gender and household wealth. Contact numbers were higher in schools than in transport, workplaces and entertainment venues, among those who attended school. After COVID-19 pandemic restrictions were imposed in early 2020, contact numbers dropped significantly in almost all groups, with the size of decline being more substantial in groups which started off with more contacts.

With respect to occupation, reported contact numbers pre-lockdown were significantly higher for students, manual labourers and service workers pre-lockdown compared to homemakers and the unemployed. When it came to lockdown reductions, these were most substantial for students and teachers, informal workers and for those living on agricultural estates. When compared to other findings in the literature, Dobreva et al (2022) note in their study across 19 African Union member states that informal workers are contact intensive – but equally do not disaggregate contacts by occupation type in the way this study does. Though they appear to cover data for Zimbabwe too, their sample is much more limited to 1,284 respondents over two sites(Dobreva *et al*., 2022). Meanwhile in Europe and North America, existing studies have documented that education, healthcare workers and customer facing/service roles tended to be high-contact and slower to reduce contacts. This appears consistent with this paper’s findings in the pre-lockdown era but not for the reductions. This paper also sheds light on how LMIC contact patterns can thus differ(Beale *et al*., 2022; Kiti *et al*., 2023).

In addition to occupation, the research shows a range of factors associated with pre-lockdown contact numbers. Some of these have been examined previously, even in LMIC settings. For example, men reported more contacts than women, which is in line with global LMIC evidence from seven contact surveys (Mousa *et al*., 2021) when accounting for age. Potentially this may be associated with occupation, given that 68% of those classified as homemakers or not employed were female versus 32% male. Informal traders were also predominantly women (73%) while the high contact occupation of manual labourers was predominantly men (71%). Teachers and students were more evenly distributed between genders (52% female) but were high contact also. In a 2013 contact survey from the same site however, the disparity in contacts between genders is less marked pointing to some change in social dynamics in this period (Melegaro *et al*., 2017). Similarly, the higher contact counts in younger age-groups is consistent with other LMIC data in southern Africa and beyond (McCreesh *et al*., 2021; Mousa *et al*., 2021). Several of the associations identified are however novel. (Quaife, van Zandvoort, *et al*., 2020). These include having higher household wealth, or living in an urban area, being associated with more contacts.

With regards to the association between contacts and wealth, this may be associated with the more rural nature of the study sites. We might speculate that higher wealth individuals in rural areas may be more able to travel and thus choose to interact with more people than those in informal urban settlements in the Kenya study. That this study found the marginal reduction in contacts to be smaller for individuals in poorer households is partially reflective of their lower number of contacts as a starting point. However there is also qualitative evidence that lower income communities in Zimbabwe found it difficult to engage in social distancing during the pandemic due to the need to earn a daily income for subsistence (Mackworth-Young *et al*., 2021). This study also finds an association between respondents with more contacts and those living in an urban area. There are also few studies that have examined the relationship between contacts and urbanity (Mousa *et al*., 2021), and despite seeming like a logical association, studies in Kenya show that rural respondents had more contacts than their urban counterparts (Kiti *et al*., 2016; Del Fava *et al*., 2021), warranting further sociological investigation. In the Zimbabwean context, that urban residents have more contacts may have important implications for infectious disease spread and associated control measures. In terms of occupation, working in manual labour or as a student, or as a teacher or in the service sector are areas that have not necessarily been highlighted in previous studies that have focused on age based mixing (Mousa *et al*., 2021).

COVID-19 lockdown measures led to a significant decrease (33% on medians overall) in numbers of reported contacts. This decrease in contact numbers was heterogeneous within the study population. The fall was proportionally greatest for high-contact individuals and for groups that previously had the highest number of contacts. This is positive in the sense that those most at risk of spreading infection were those who most drastically curtailed their activities. For example, the greatest marginal reductions in contact numbers were in males by gender and those aged 30-44 by age group. Distinctions by age are consistent with studies in South Africa (McCreesh *et al*., 2021), Kenya (Del Fava *et al*., 2021), Malawi (Thindwa *et al*., 2021), Uganda (le Polain de Waroux *et al*., 2018) and many other LMICs (Mousa *et al*., 2021), where younger adults both had the largest number of contacts in the population before lockdown and experienced the largest drop proportional to the over 65s. Other significant drops reported in this study were experienced especially in students, teachers and informal traders by occupation; urban and agricultural estates by site. The distinctions by site type vary in other studies. For example, in urban Malawi, only small changes were found in contacts in response to restrictions (Thindwa *et al*., 2021), whilst, in informal settlements in Nairobi, Kenya, a 62% reduction in physical contacts and 63%-67% reductions in non-physical contacts were found (Quaife, van Zandvoort, *et al*., 2020).

In this study, household wealth groups saw similar reductions, which has not been explored in other studies reviewed here. By contact place, the biggest drop was reported at school – reflecting the total cessation of in-person teaching in Zimbabwe which took place between 22^nd^ March – 4^th^ June 2021 during this study’s data collection period. There were drops in contacts in the workplace and on transport as well as in entertainment venues. Some occupations e.g. informal workers and traders saw marginal reductions of between 11 and 13 contacts and constitute a total of 19% of the population. This study highlights the importance of pulling out these distinctions and looking at the impact that differentiated interventions might make to limiting the spread of COVID-19 in Zimbabwe by factors such as occupation, or school attendance, as was conducted elsewhere (Chao *et al*., 2020; Panovska-Griffiths *et al*., 2020; Chung *et al*., 2023; Domenico *et al*., no date).

Existing studies that have examined the relationship between occupation and transmission risk of COVID-19 have been in Europe and North America - such as in the Netherlands (De Gier *et al*., 2020), Norway (Magnusson *et al*., 2020), Romania (Hâncean *et al*., 2022) and a study that looked at relative proximity in different workplaces in California, USA(Huang *et al*., 2020). This paper has contributed to understanding differences in social contacts in workplaces in a LMIC context – rather than these High Income Country contexts, that can be used to inform context specific infectious disease models. Workplaces in LMICs are known to differ due to a range of factors, including higher levels of informality.

Social mixing data is essential for building mathematical models that seek to replicate the transmission dynamics of an infectious disease outbreak. Outcomes can differ when considering different contact levels and measures to control for them (Walker *et al*., 2020) and many countries lack any local reference data (Mousa *et al*., 2021). This study contributes to understanding the impact of applying non-pharmaceutical interventions such as movement restrictions and bans on gatherings, by adding to the knowledge on the number of contacts that typically would happen in those contact places and by occupation type. Due to the lack of information on how COVID-19 lockdown restrictions actually impacted peoples contacts in many countries; even the self-reported estimates of the changes presented here can give us a clearer idea of how impactful they were for different population subgroups.

A recommendation for future work to enhance the richness of the data presented here would be to conduct further qualitative analysis on the nature or duration of reported interactions, or how they could contribute to epidemic spread. A richer understanding of how social contact leads to the spread of infection could be gained from integrating local knowledge based on theories of human behaviour (Buckee, Noor and Sattenspiel, 2021), as well as and a deeper analysis of the degree of mixing assortativity between and within different population subgroups. The latter would require additional data on, e.g., kin structures, mixing contexts (religious and social gatherings, marketplaces, social visiting) and seasonal behaviours (labour migration, seasonal agriculture). Behavioural survey data could also complement this study’s findings and to provide a better understanding of how contact counts could be reduced. Some prior rapid qualitative research carried out in Zimbabwe in the early lockdown period (March and April 2020) revealed widespread fear, due to uncertainty over credible information, and reported difficulties for some communities to comply with social distancing measures when households needed to access water and food supplies for daily subsistence (Mackworth-Young *et al*., 2021). Further qualitative research would help to shed light on whether the reported high reductions were due to government policy versus people deciding not mix due to their own fear of getting infected. Forthcoming work conducted by the same authors has collected semi-structured interview data on a sub-sample of the same population’s responses to the government’s movement restrictions which may shed more light on this question.

### Limitations

Despite its value, the study has some limitations. First, this study’s social contact distributions seem to contain more noise than data collected from the same site a decade earlier. In a social contact study in Manicaland in 2013, participants reported slightly lower median weekday contact numbers of 9 (6-14) vs 12 in our data with a much wider IQR(4-47), and fewer extreme counts (Melegaro *et al*., 2017). These differences may reflect the nature of this study’s survey questions, which asked for the number of contacts within two meters on a ‘typical day’ rather than using an activity diary. This relatively abstract question may have led to contact inflation (as was evident from the ‘other’ contacts removed from the calculation – see Text S3 and Table S9 for details). Alternatively, however, differences may also reflect the wider range of sites included in the current study, including towns, tea estates and other locations with high potential for congregate events. The authors suggest that the greater value of this study’s data lies in the patterns of contact and contact reduction, rather than the absolute numbers reported.

Relatedly, the fact that the contact numbers were self-reported may lead them to be subject to recall bias or desirability bias. As participants were asked about their present contacts prior to their pre-lockdown contacts, there would be less likelihood of under-reporting their present contact numbers. However, there could still be a tendency for participants to have over-inflating the contacts they had prior to the restrictions to demonstrate how they had changed their behaviour. Some other studies have attempted to use sensors or passes to measures contact numbers in institutional spaces such as hospitals(Wilson-Aggarwal *et al*., 2023) or in schools (Kiti, Moses Chapa, 2019). However, when measuring contacts across a whole community, the use of sensors through e.g. Bluetooth devices introduces other inaccuracies(Leith and Farrell, 2020). Second, generalizing the findings presented here to other populations requires consideration of differences in wealth and occupation patterns, as well as the types of lockdowns imposed. The findings of this study are likely to be most applicable in rural and small-town settings in other Sub-Saharan African countries. On the other hand, the lack of generalizability of the data is arguably offset by the reasonably representative selection of site types being included in the study.

## Conclusions

The number of social contacts that an individual has can greatly affect their likelihood of spreading many infectious diseases, and movement restrictions affect population subgroups to differing degrees. While several social contact studies have shown how contact patterns changed during blanket Covid-19 lockdowns, this study shows that these patterns were also patterned by occupation as well as demographic characteristics, in a setting with very limited formal employment. The study highlights that movement restriction policies targeted to those groups with the highest non-pandemic contact levels could reduce disease spread and mitigate economic harm, something especially important in LMICs with large informal economies. Future deeper exploration of the mechanisms through which wealth and occupation are associated with the impact of lockdown regulations should inform the design more targeted pandemic responses. The study also sheds light on what remains unknown about the reasons for why people may choose to reduce their social contacts during an outbreak and thus highlights the need for further qualitative research to understand the behavioural motivations for contact reductions. Overall, using the data this study has collected show that contacts can differ by other socio-economic factors beyond age, including occupation and thus policy prescriptions may consider how social safety nets should be tailored when restricting the capacity of some occupations to earn a living.

## Data Availability

Data can be made available on request to the Manicaland Centre for Public Health Research. A data request form is available at http://www.manicalandhivproject.org/manicaland-data.html

## List of Abbreviations

MRC-Z: Medical Research Council of Zimbabwe
RSV: Respiratory Syncytial Virus
SARS: Severe Acute Respiratory Syndrome
HICs: High Income Countries
SSA: Sub-Saharan Africa
LMICs: Low and Middle Income Countries
IRR: Incidence Rate Ratio
CI: Confidence Interval
COVID-19: Coronavirus Disease 2019
AIDS: Acquired Immunodeficiency Syndrome
CDC: Centers for Disease Control and Prevention
FCDO: Foreign, Commonwealth and Development Office
HIV: Human Immunodeficiency Virus
IPUMS: Integrated Public Use Microdata Series
IQR: Interquartile Range
LD: Lockdown
MRC: Medical Research Council
NFID: National Foundation for Infectious Diseases
PLD: Pre-lockdown
SARS-CoV-2: Severe Acute Respiratory Syndrome Coronavirus 2
TB: Tuberculosis
UCL: University College London
UNDP: United Nations Development Programme
WHO: World Health Organization

## Declarations

### Authors’ contributions

SA - Conceptualization, methodology, formal analysis, investigation, data curation, writing – original draft and editing, visualization

CN – Project administration

RMa – Project administration

LM – Data curation, project administration

RMo – Data curation

BM – Resources, Supervision

AM – Conceptualization, methodology

OM - Resources, Supervision

HF - Supervision SW – Supervision

GH – Supervision, conceptualization, methodology, resources, writing – reviewing and editing, funding acquisition

SG – Supervision, conceptualization, methodology, resources, writing – reviewing and editing, funding acquisition

All authors approved the final version for submission and agree to be accountable for the integrity of the work

## Acknowledgements

We are very grateful to the Manicaland Centre for Public Health Research team, particularly Phyllis Mandizvidza, Freedom Dzamatira, Tawanda Dandirai and Blessing Tsenesa for their work on the data collection, and to study participants in Manicaland, east Zimbabwe, for their participation in and contribution to this work; The write up also received valuable inputs from Filippo Trentini.

This study is dedicated to memory of Dr. Constance Nyamukapa who contributed to the development of this research before her passing. Her commitment to the Manicaland Centre for Public Health Research and generosity of spirit as a colleague and friend made the fieldwork possible. We hope this work honours her contribution.

## Declaration of generative AI and AI-assisted technologies in the manuscript preparation process

During the preparation of this work the authors used Claude Code (Anthropic) in order to carry out a review of the statistical analysis code prior to submission, to verify that the tables reported in this manuscript and its supplementary information were consistent with the output of the underlying replicable analysis files, and to draft the graphical abstract. The graphical abstract is a data visualisation derived directly from the study data by a reproducible analysis script. After using this tool, the authors reviewed and edited the content as needed and take full responsibility for the content of the published article.

## Supporting Information

**Fig S1** Trends in reported national COVID-19 infections and deaths, 2020 to 2022.

**Table S1** Details on lockdown periods and government responses in response to the COVID-19 pandemic in Zimbabwe 2020-2021.

**Table S2** Study sites of Manicaland Survey.

**Table S3** Extract from Individual Round 8 Questionnaire with questions analysed.

**Text S1** Ethical Approvals – Medical Research Council of Zimbabwe, Imperial College London and University College London

**Text S2** Consent forms as approved by MRCZ and RCZ

**Text S3** Supplementary analysis on Data Pre-Processing and Outlier Management.

**Table S4** Comparison of ordinary, cluster-robust and mixed-effects negative binomial models for pre-lockdown contacts.

**Table S5** Raw differences in contact numbers by place between lockdown and non-lockdown.

**Table S6** Margins differences in contacts non-lockdown vs. lockdown (population level).

**Table S7** Regression on the difference — correlates of changes over all places and age groups.

**Table S8** Nested Regressions on determinants of differences by place between lockdown and non-lockdown contact rates (population level — except school, at place level).

**Table S9** Descriptive statistics for pre-lockdown and lockdown contacts with ‘other’ contacts included (sensitivity analysis).

## Notes

### Competing Interest Statement

SG declares shareholdings in pharmaceutical companies (GlaxoSmithKline and AstraZeneca).

### Clinical Trial

NA

### Author Declarations

The Manicaland Survey was approved by the Imperial College Research Ethics Committee (20IC6436) and the Medical Research Council of Zimbabwe (MRCZ/A/2703). This analysis was approved by the University College London Research Ethics Committee (project ID 20527/001) All participants provided audio-recorded (for telephone interviews) or written (for face-to-face interviews) informed consent prior to participation.

